# Efficacy, safety and predictors of response to electroconvulsive therapy in Lewy body disease

**DOI:** 10.64898/2025.12.02.25341437

**Authors:** Matasaburo Kobayashi, Yoshiyuki Nishio, Yuto Satake, Hideki Kanemoto, Kumiko Utsumi, Manabu Ikeda

**Affiliations:** Department of Psychiatry, The University of Osaka Graduate School of Medicine, Suita, Japan; Department of Behavioural Neurology and Neuropsychiatry, The University of Osaka United Graduate School of Child Development, Suita, Japan; Department of Psychiatry, Asakayama General Hospital, Sakai, Japan, The University of Osaka Graduate School of Medicine, Suita, Japan; Health and Counseling Center, The University of Osaka, Toyonaka, Japan; Hakuyukai Takikawa Mental Clinic, Takikawa, Japan

## Abstract

**Background:** Severe, treatment-resistant psychiatric symptoms are common in Lewy body disease (LBD). As pharmacotherapy is often limited by poor efficacy and adverse effects, alternative treatments are needed. While electroconvulsive therapy (ECT) has shown promise, systematic data on its use in LBD remain scarce.

**Aims:** To evaluate the efficacy and safety of ECT in patients with LBD and identify predictors of a favorable response.

**Method:** We compared 40 patients with LBD to 33 with schizophrenia and 24 with affective disorders who received ECT between 2012 and 2023. The primary outcome was short-term efficacy, measured by the Clinical Global Impressions-Improvement (CGI-I) scale. Long-term outcomes were assessed via 6-month and 2-year readmission rates. An ordinal logistic regression analysis was used to identify clinical predictors of response, including target symptoms (psychosis, catatonia, and depression), in the LBD group.

**Results:** Short-term efficacy in the LBD group was comparable to that in the schizophrenia group but lower than in the affective disorders group (median CGI-I: 2 vs. 1, p < 0.05). Within the LBD group, the regression analysis revealed that psychosis and catatonia were significant predictors of a favorable response (p = 0.044). While 6-month readmission rates were similar across groups, the 2-year rate was highest among LBD patients (61.5%). ECT was well-tolerated, with no serious adverse events and transient amnesia as the most common side effect.

**Conclusions:** ECT is an effective and safe treatment for severe psychiatric symptoms in LBD, particularly for patients with psychosis or catatonia. Although its short-term efficacy may be less pronounced than in affective disorders, it represents a valuable therapeutic alternative when pharmacotherapy is challenging. Further research is warranted to confirm long-term benefits and optimize patient selection.

## Introduction

Lewy body disease (LBD), which encompasses Parkinson’s disease (PD) and dementia with Lewy bodies (DLB), is a leading cause of movement and cognitive disorders in older adults. PD accounts for two-thirds of parkinsonism in people aged 55 years and over,^1^ whereas DLB accounts for 7.5% or more of all dementias in people older than 65 years.^2^ LBD is increasingly recognised as presenting with diverse psychiatric symptoms, including depression, anxiety, psychosis and impulse control disorders.^3^ The onset of these psychiatric symptoms is highly variable, ranging from phases preceding the full development of motor or cognitive syndromes to later stages long after the diagnosis of PD or DLB is established.^4,5^ LBD-associated psychiatric symptoms are highly prevalent and significantly affect the quality of life of patients and carers. Psychosis and depression are observed in the majority of patients with DLB and are listed as the core and suggestive clinical features in the current diagnostic criteria.^6^ Point prevalence estimate for depression and psychosis in PD reach up to 35% and 25-40%, respectively, and more than the half of patients with PD eventually develop psychosis.^3^ Previous studies have demonstrated that psychiatric symptoms pose a greater carer burden in DLB than in Alzheimer’s disease.^7,8^ Similarly, depression and psychosis are common and are associated with higher mortality and admission rates to care facilities in PD.^9,10^

Psychiatric symptoms represent a profound clinical challenge in both PD and DLB, often dictating patient prognosis more significantly than motor dysfunction or cognitive impairment.^11,12^ While depression and psychosis are prevalent in LBD, profound agitation and aggression frequently complicate the clinical picture, often in association with hallucinations and delusions.^13^ Crucially, psychomotor alterations can escalate to catatonia, which is not merely a behavioural disturbance but a life-threatening syndrome associated with high mortality due to complications such as deep vein thrombosis, pulmonary embolism, and malnutrition.^14^ Consequently, immediate symptomatic resolution is imperative. The therapeutic management of these neuropsychiatric syndromes is fraught with complexity, presenting clinicians with a profound ‘therapeutic dilemma’: dopaminergic agents used to treat motor symptoms may exacerbate psychosis, while dopamine antagonists used to treat psychosis often worsen motor function, thereby limiting the utility of conventional pharmacotherapy. Notably, patients with LBD exhibit a distinct hypersensitivity to antipsychotic medications, which precludes dose escalation to therapeutic levels.^15^ Furthermore, evidence for the efficacy of commonly used agents like quetiapine remains equivocal in randomised trials,^16^ and novel agents such as pimavanserin carry specific regulatory warnings regarding their use in dementia populations.^11^ In this context of therapeutic deadlock, electroconvulsive therapy (ECT) has emerged as a vital treatment option for drug-resistant psychiatric symptoms in LBD, particularly when pharmacotherapy is ineffective or contraindicated.^12^

Recent systematic reviews indicate that ECT demonstrates robust efficacy for depression, catatonia, and psychosis within the LBD spectrum, with a safety profile characterized primarily by transient rather than persistent delirium.^12^ Despite this clinical utility, the current evidence base relies heavily on case series and retrospective reports, and systematic investigations regarding its efficacy, tolerability, and specific target symptoms remain scarce. This study retrospectively examined clinical data accumulated over a 10-year period at a single institution to investigate the overall efficacy of ECT for patients with LBD, and to identify predictors of a positive response. To contextualize the findings, patients with affective disorders and schizophrenia, psychiatric disorders for which substantial evidence on ECT exists, were used as comparison groups.

## Methods

### 1. Ethical considerations and study design

The authors assert that all procedures contributing to this work complied with the ethical standards of the relevant national and institutional committees on human experimentation and with the Declaration of Helsinki of 1975, as revised in 2013. All procedures involving human participants were approved by the Regional Ethics Committee of Sunagawa City Hospital and The University of Osaka Hospital Ethical Review Board. This study was conducted as a retrospective observational study using existing clinical data. According to the institutional review board policy, individual informed consent was waived because the study involved no more than minimal risk to the participants. Instead, an opt-out approach was employed, and information regarding the study was publicly disclosed on the hospital website to provide patients with the opportunity to refuse participation. All participant data were anonymized before analysis to ensure patient confidentiality.

### 2. Participants

We identified potential participants from patients who received their first ECT at Sunagawa City Hospital between January 2012 and December 2023. LBD diagnoses were subsequently established based on: 1) fulfilment of the criteria for possible/probable DLB,^6^ PD,^17^ or PD dementia,^18^ or 2) the presence of an LBD biomarker when neither the core clinical features nor the evidence of dementia was present, a state we defined as prodromal LBD.

We recruited two distinct comparison groups from the same patient pool: one consisting of patients with affective disorders (major depressive disorders and bipolar disorder) and another with schizophrenia. Diagnoses for these groups were made according to the DSM-IV-TR and DSM-5. If a patient tested positive for an LBD biomarker, they were assigned to the LBD group regardless of their psychiatric diagnosis.

The exclusion criteria were as follows: 1) unspecified target symptoms for ECT 2) a comorbid stroke or other neurodegenerative diseases or psychiatric disorders; and 3) premature termination of ECT for reasons unrelated to treatment efficacy or side effects.

### 3. ECT procedure

ECT was administered using a brief-pulse square-wave device (Thymatron™ System IV; Somatics, LLC, Lake Bluff, IL, USA). Bilateral frontotemporal electrode placement was used for all patients. Patients typically received 6–12 sessions of ECT. However, treatment was terminated early if clinicians deemed it ineffective, or extended if further improvement was anticipated. Stimulus intensity was determined via the age-based method (approximate age in years % setting) for the first session and subsequently titrated based on seizure quality. A pulse width of 0.5 ms was used. Anesthesia was induced with propofol and succinylcholine, following pre-oxygenation via a bag-valve mask. Seizure adequacy was monitored using electroencephalography (EEG), with a target seizure duration of at least 20 seconds.

### 4. LBD biomarkers

#### ∂ Dopamine transporter imaging

A subset of patients underwent^123^I-FP-CIT dopamine transporter (DaT) SPECT. Three raters (YN, YS, and HK) visually assessed the blinded SPECT images according to the previously proposed grading criteria: Normal; Grade 1, asymmetric uptake with normal putamen uptake in one hemisphere and with a more marked reduction in the contralateral putamen; Grade 2, significant bilateral reduction in putamen uptake with activity confined to the caudate nuclei; Grade 3, virtually absent uptake bilaterally affecting both putamen and caudate nuclei.^19^ A rating of Grade 1–3 by two or more raters was considered indicative of a positive LBD biomarker. None of the patients who underwent DaT-SPECT were taking medications that potentially affect DaT uptake.^20^

#### ∂ Cardiac MIBG imaging

To assess myocardial sympathetic innervation, a subset of patients underwent cardiac^123^ I-MIBG imaging. A delayed heart-to-mediastinum ratio <2.2 was considered positive.^21^ Two patients who were taking tricyclic antidepressants at the time of MIBG myocardial scintigraphy were excluded. The other patients were not taking medications that potentially affect MIBG uptake.^22^

### 5. Clinical assessments

To contextualize the efficacy of ECT for LBD, we compared the efficacy of ECT in the LBD group with that in the affective disorder and schizophrenia groups.

The short-term efficacy of ECT was assessed using the Clinical Global Impressions-Severity/Improvement (CGI-S/I).^23^ Three board-certified neuropsychiatrists (YN, YS, and HK), blinded to patient diagnoses, sex and age, evaluated the CGI. The CGI-S/I ratings were exclusively based on summarized excerpts from medical records, which included patient behaviour, dietary intake and sleeping patterns during the weeks immediately preceding and following the ECT course. The median score from the three raters was used for subsequent statistical analysis. Long-term ECT efficacy was assessed by the patient re-admission rate at six months and two years after completion of the ECT course.

We classified the target psychiatric symptoms for ECT into the three categories: psychosis, catatonia and depression. These categories were not mutually exclusive. Side effects were identified through a retrospective review of medical records during the ECT course and for one week following its completion.

### 6. Statistical analysis

Comparisons of demographic and clinical characteristics were made among the three groups (LBD, affective disorders and schizophrenia) using one-way analysis of variance for continuous variables (ANOVA) (age, MMSE scores, ECT sessions), the Fisher’s exact test for the categorical variables (sex, targeted symptoms, readmission within a certain period, adverse events), and the Kruskal-Wallis test for ordinal variables, followed by the Dunn–Bonferroni test for post-hoc pairwise comparisons when appropriate (CGI scores).

To identify predictors of a favourable ECT response in the LBD group, an ordinal logistic regression analysis was performed with the CGI-I as the outcome variable and with age, sex, and target symptom categories (psychosis, catatonia, and depression) as explanatory variables.

A p-value <0.05 was considered statistically significant. These analyses were performed using JMP Pro 17.1.0. (JMP Statistical Discovery LLC., Cary, NC, USA).

## Results

### 1. Participants and demographics

Flow diagram of participant selection is shown in **Figure 1**. Of the 119 patients initially assessed, 97 were included in the final analysis. Among the 97 patients, 40 were diagnosed with LBD (21 with probable DLB, 7 with possible DLB, 7 with prodromal DLB, 1 with PD and 4 with PDD), 33 with schizophrenia, and 24 with affective disorders (18 with major depressive disorder and 6 with bipolar depression). Twenty-two patients were excluded for the following reasons: 20 met the exclusion criteria and 2 were taking tricyclic antidepressants at the time of MIBG imaging.

**Figure 1.**
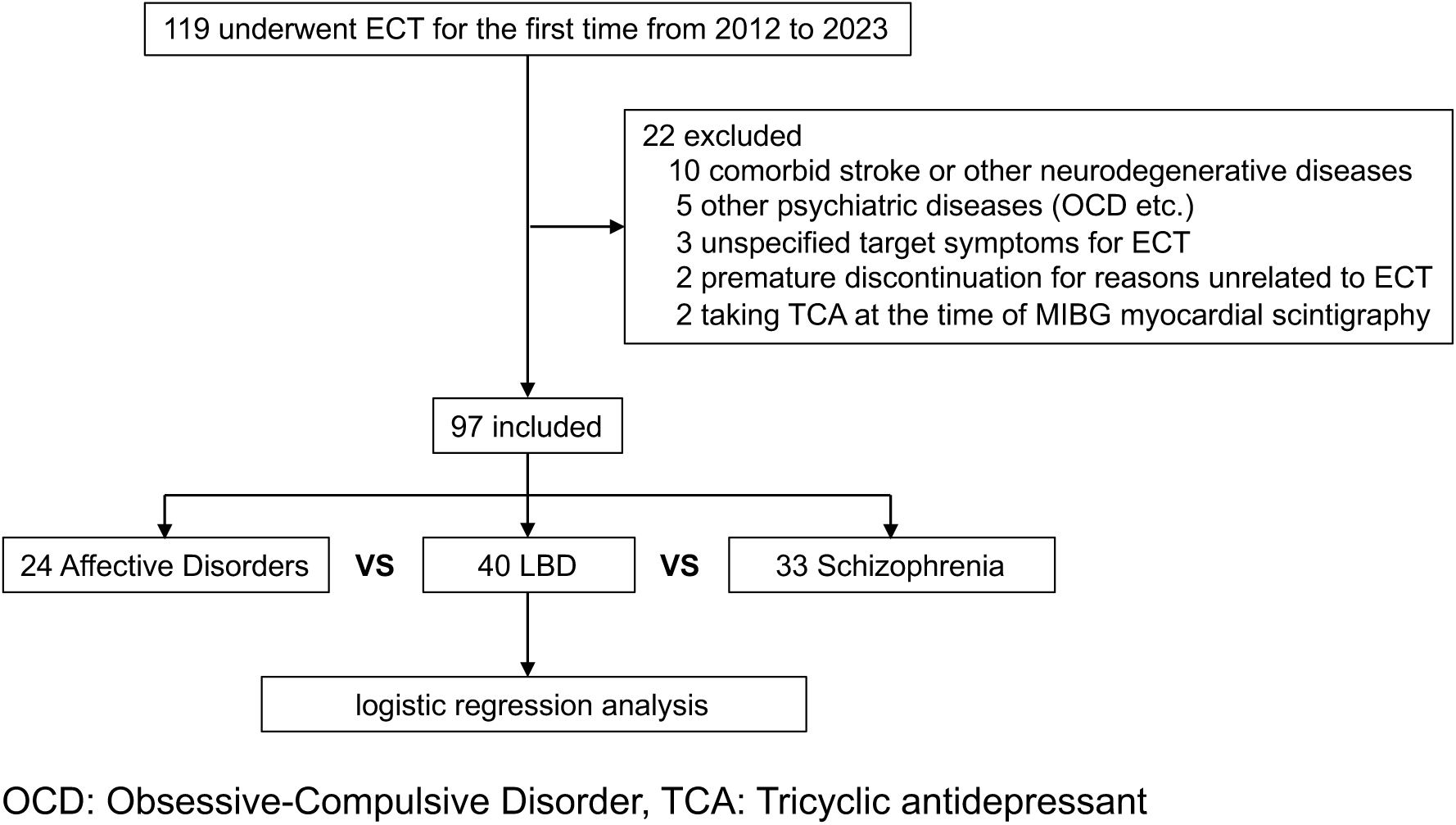
Flow diagram of participant selection.

Demographic and clinical characteristics of the participants are summarised in **Table 1**. The DLB group had a significantly higher mean age than the schizophrenia and affective disorders groups (one-way ANOVA, p < 0.001). Among the 40 patients with LBD, 24 (60.0%) had visual hallucinations, 22 (55.0%) had cognitive fluctuation, 19 (47.5%) had parkinsonism, and 11 (27.5%) had rapid eye movement sleep behaviour disorder (RBD).

**Table 1.** Demographic and clinical characteristics of participants.

|  | LBD<br>(N=40) | Schizophrenia<br>(N=33) | Affective disorders<br>(N=24) | P-value |
| --- | --- | --- | --- | --- |
| Age, mean (SD) | 75.1 (7.9) | 47.2 (13.1) | 64.7 (10.7) | **<0.001** |
| Female, n (%) | 27 (67.5%) | 20 (60.6%) | 18 (75.0%) | 0.52 |
| Baseline MMSE, mean (SD) | 20.7 (4.9, N=27) | 24.7 (4.9, N=3) | 17.5 (10.6, N=2) | 0.458 |
| LBD biomarker positivity |  |  |  |  |
| DaT scan, n (%) | 18/35 (51.4%) | 0/4 (0%) | 0/13 (0%) |  |
| MIBG, n (%) | 8/15 (53.3%) | 0/1 (0%) | 0/3 (0%) |  |
| DLB Core Symptoms |  |  |  |  |
| Visual hallucination, n (%) | 24 (60.0%) | - | - |  |
| Cognitive fluctuation, n (%) | 22 (55.0%) | - | - |  |
| Parkinsonism, n (%) | 19 (47.5%) | - | - |  |
| RBD, n (%) | 11 (27.5%) | - | - |  |

MMSE: Mini-Mental State Examination, DaT scan: dopamine transporter scintigraphy, MIBG: MIBG myocardial scintigraphy, RBD: REM sleep behaviour disorder

### 2. Treatment characteristics and outcomes of ECT

Treatment characteristics and outcomes of ECT are summarised in **Table 2**. There were no significant differences in pre-ECT clinical severity, as measured by the CGI-S, among the LBD, schizophrenia, and affective disorders groups. The mean number of ECT sessions was 8.9 (2.3) in the LBD group and did not significantly differ from those in the other two groups. In the LBD group, the most frequent target symptoms of ECT was psychosis (70.0%), followed by depression (50.0%) and catatonia (30.0%). The majority of patients with schizophrenia received ECT for psychosis (97.0%), whereas most patients with affective disorders received treatment for depression (95.8%).

**Table 2.** Treatment Characteristics and outcomes of ECT.

|  | LBD<br>(N=40) | Schizophrenia<br>(N=33) | Affective disorders<br>(N=24) | P-value |
| --- | --- | --- | --- | --- |
| ECT sessions, mean (SD) | 8.9 (2.3) | 8.6 (2.6) | 8.5 (2.0) | 0.763 |
| Targeted symptoms |  |  |  |  |
| psychosis, n (%) | 28 (70.0%) | 32 (97.0%) | 6 (25.0%) | **<0.001** |
| catatonia, n (%) | 12 (30.0%) | 13 (39.4%) | 11 (45.8%) | 0.423 |
| depression, n (%) | 20 (50.0%) | 7 (21.2%) | 23 (95.8%) | **<0.001** |
| CGI |  |  |  |  |
| Baseline CGI-S, median [IQR] | 7 [6-7] | 7 [6-7] | 7 [6-7] | 0.477 |
| Post-ECT CGI-S, median [IQR] | 3 [2-4] <sup>a*</sup> | 4 [2-4] <sup>a</sup> | 2 [1-2] <sup>b,c</sup> | **<0.001** |
| CGI-I, median [IQR] | 2 [1-2] <sup>a</sup> | 2 [1-2] <sup>a</sup> | 1 [1-2] <sup>b,c</sup> | **0.007** |
| Readmission |  |  |  |  |
| 6 months, n (%) | 9/31 (29.0%) | 6/28 (21.4%) | 5/22 (22.7%) | 0.771 |
| 2 years, n (%) | 16/26 (61.5%) | 10/25 (40.0%) | 7/20 (35.0%) | 0.146 |

ECT: Electroconvulsive therapy, CGI-S/I: Clinical Global Impressions-Severity/Improvement
\*Superscript letters denote statistically significant differences identified in the post-hoc analysis. a: vs Affective disorders, b: vs LBD, c: vs Schizophrenia

Catatonia was present in 39.4% of the schizophrenia group and 45.8% of the affective disorders group. While the frequencies of psychosis and depression differed significantly among the three groups (Fisher’s exact test, p < 0.001 and p < 0.001, respectively), the frequency of catatonia did not (p = 0.423).

Clinical improvement at one week post-ECT differed significantly among the three groups, with the affective disorders group showing significantly better improvement than both the LBD and schizophrenia groups (median [IQR] CGI-I score was 2 [1-2] in the LBD group, 2 [1-2] in the schizophrenia group, and 1 [1-2] in the affective disorders group). There was no significant difference in clinical improvement between the LBD and schizophrenia groups.

The patient re-admission rates at six months post-ECT did not differ significantly among the three groups (29.0%, 21.4%, and 22.7% for the LBD, schizophrenia, and affective disorders groups, respectively). The two-year re-admission rate was highest in the LBD group (61.5%), followed by the schizophrenia group (40.0%) and the affective disorders group (35.0%), although these differences did not reach statistical significance. The number of patients lost to follow-up at six months/two years was 9/14 for LBD, 5/8 for schizophrenia, and 2/4 for affective disorders. Reasons for loss to follow-up included nursing home admission and transfers to other hospitals, and unknown causes.

### 3. Predictors of ECT response in LBD

The results of an ordinal logistic regression analysis of predictors for ECT response in LBD are summarised in **Table 3**. The presence of catatonia and psychosis was significantly associated with a more favourable response to ECT in the LBD group (log-likelihood ratio test for the overall model: p = 0.0436).

**Table 3.** Ordinal logistic regression analysis of predictors for ECT response in LBD.

| Independent variables | $\beta$ | SE | OR (95%CI) | P-value |
| --- | --- | --- | --- | --- |
| age | -0.056 | 0.047 | 0.791 (0.399, 1.558) | 0.23 |
| sex (female:1, male: 0) | -0.235 | 0.347 | 0.945 (0.857, 1.306) | 0.495 |
| psychosis (yes:1, no: 0) | -0.906 | 0.426 | 0.405 (0.164, 0.930) | **0.033** |
| catatonia (yes:1, no: 0) | -0.827 | 0.383 | 0.438 (0.198, 0.890) | **0.022** |
| depression (yes:1, no: 0) | -0.46 | 0.379 | 0.632 (0.300, 1.291) | 0.208 |

$\beta$ : regression coefficient, SE: standard error, OR: odds ratio, 95%CI: 95% Confidence Interval, LR test: likelihood ratio test

### 4. Safety and adverse events

A summary of adverse events is provided in **Table 4**. The most frequently reported adverse event was transient amnesia, occurring in 7 patients in the LBD group, 10 in the schizophrenia group, and 11 in the affective disorders group. The incidence of amnesia differed significantly among the three groups (p = 0.002), with the LBD group showing the lowest incidence. Post-ECT delirium was observed in one patient in the LBD group and one in the affective disorders group, with no significant between-group difference (p = 0.206). One patient in the schizophrenia group developed status epilepticus.

**Table 4.** Adverse events associated with ECT.

|  | LBD | Schizophrenia | Affective disorders | P-value |
| --- | --- | --- | --- | --- |
|  | (N=40) | (N=33) | (N=24) |  |
| Delirium, n (%) | 1 (2.5%) | 0 (0%) | 1 (4.2%) | 0.206 |
| Amnesia, n (%) | 7 (17.5%) | 10 (30.3%) | 11 (45.8%) | **0.002** |
| Others*, n (%) | 1 (2.5%) | 1 (3.0%) | 1 (4.2%) | - |

\*Others included gallstone pancreatitis in a patient with LBD, status epilepticus in a patient with schizophrenia, and cholecystitis in a patient with an affective disorder.

## Discussion

This study provides a long-term retrospective analysis of ECT in patients with LBD, comparing outcomes with primary psychiatric disorders. Our results demonstrate that ECT is a viable and safe therapeutic option for severe, treatment-resistant psychiatric symptoms in LBD. While the degree of improvement was less pronounced than in primary affective disorders, it was comparable to schizophrenia, offering a crucial intervention for patients who are often refractory to pharmacotherapy.

## Efficacy and Safety Profile

In the present study, ECT yielded a clinically meaningful improvement in psychiatric symptoms associated with LBD, achieving a median CGI-I score of 2 (“Much Improved”). This degree of efficacy was comparable to that observed in the schizophrenia group but was significantly less pronounced than in the affective disorders group, where patients frequently achieved complete remission. These findings are consistent with a recent systematic review and multicentre survey, which reported that ECT is generally effective for depression, catatonia, and psychosis in patients with DLB, although the magnitude of response can vary compared to primary psychiatric conditions.^12,24^ Given that the use of antipsychotic medications is often restricted in LBD due to the risk of severe sensitivity reactions and motor deterioration, ECT represents a vital alternative strategy for managing severe, drug-resistant symptoms.

Regarding safety, the incidence of post-ECT delirium in our LBD cohort was 2.5%, which is notably lower than the rates of 16.1% to 24% reported in previous studies.^12,24^ Since this study, like previous ones, was retrospective, this discrepancy is likely attributable to differences in the criteria for documenting adverse events or the threshold for including mild, transient confusion, rather than differences in patient selection. It is possible that minor episodes of delirium were underreported in the medical records used for this analysis. However, consistent with reports on ECT in schizophrenia and major depression,^25^ we observed no serious, irreversible adverse events or permanent cognitive decline. This suggests that while the risk of delirium exists, ECT can be administered with an acceptable safety margin in this population when appropriate monitoring is in place.

## Psychomotor Symptoms and Dopaminergic Mechanisms as Transdiagnostic Predictors

A key finding of this study is that psychosis and catatonia, but not depression, predicted a favorable ECT response within the LBD group. This parallels findings in major depressive disorder and schizophrenia, where the presence of psychotic features or catatonia is consistently associated with superior ECT outcomes.^25–27^ Ideally, depression would also be a robust predictor, yet its lack of significance in our model highlights the heterogeneity of “depression” in the context of LBD. These findings can be integrated through the lens of striatal dopamine dysfunction. It has been proposed that reduced striatal dopamine transmission serves as a transdiagnostic substrate for “psychomotor alterations,” a concept encompassing the psychomotor retardation of melancholia, the immobility of catatonia, and the motor deficits of Parkinsonism.^28^ This hypothesis is supported by recent neuroimaging evidence linking striatal dopamine dysregulation to mood instability and psychosis across different psychiatric conditions.^29^ Since ECT is known to enhance dopaminergic transmission and modulate receptor sensitivity,^30,31^ it is mechanistically well-suited to reverse a functional striatal dopaminergic insufficiency.

The differential response between ‘dopamine-responsive’ symptoms (psychosis, catatonia) and non-responsive depression warrants further scrutiny. Depression in LBD is likely multifactorial.^32^ One subtype may be driven by nigrostriatal dopaminergic degeneration, manifesting with apathy and psychomotor slowing, which theoretically responds well to dopaminergic stimulation via ECT.^28,33^ However, another subtype might be driven by cholinergic deficits, cortical atrophy, or noradrenergic dysfunction, which may be less responsive to the dopamine-enhancing effects of ECT.^32,10^ Our finding that ‘depression’ as a broad category did not predict response suggests that clinical phenotyping based on psychomotor features—rather than DSM-based diagnostic labels—may be more essential in LBD.^28^

## Clinical implications

Our findings have direct implications for clinical practice. The management of severe psychiatric symptoms in LBD often reaches an impasse where pharmacotherapy fails or causes unacceptable side effects. Our data support the earlier consideration of ECT in the treatment algorithm, particularly for patients presenting with psychosis or catatonic features. Although the remission rate may be lower than in primary depression, the goal of treatment in advanced LBD is often symptom palliation and manageability rather than complete cure. The reduction of agitation and psychosis to a level that allows for home care or reduces distress is a clinically significant outcome.

Furthermore, the safety profile observed in our study should reassure clinicians who may hesitate to prescribe ECT for older adults with neurodegenerative conditions due to fears of cognitive worsening or somatic complications.

## Limitations

This study has several limitations. First, our assessment of efficacy was limited to the short term, as the primary outcome measure, the CGI-I, was evaluated only one week after treatment completion. The long-term durability of the response could not be fully ascertained, highlighting the need for studies with extended follow-up periods. Second, because biomarker testing was not performed systematically, some patients with prodromal LBD may have been misclassified into the primary psychiatric disorder groups. Previous research indicates that a small percentage of patients with late-life depression eventually convert to LBD.^34^ Third, the higher rate of loss to follow-up in the LBD group represents a potential source of bias, as this differential attrition could have artificially inflated the observed long-term efficacy. Finally, our reliance on retrospective chart review for safety data, rather than systematic prospective monitoring, may have resulted in an underestimation of the incidence of milder adverse events.

## Conclusion

ECT is an effective and safe therapeutic strategy for severe psychiatric symptoms in LBD, particularly in cases presenting with psychosis or catatonia. The response profile suggests a transdiagnostic mechanism potentially linked to the reversal of striatal dopaminergic dysfunction. While short-term gains are significant, the progressive nature of LBD necessitates careful long-term management planning. Future prospective studies incorporating biomarkers should focus on identifying the specific biological phenotypes within LBD that yield the highest response to ECT.

## Data Availability

All data produced in the present study are available upon reasonable request to the authors.

## References

1 De Rijk MC, Breteler MMB, Graveland GA, Ott A, Grobbee DE, Van Der Meche FGA, et al. Prevalence of Parkinson’s disease in the elderly: The Rotterdam Study. Neurology 1995; 45: 2143–6.

2 Walker Z, Possin KL, Boeve BF, Aarsland D. Lewy body dementias. The Lancet 2015; 386: 1683–97.

3 Weintraub D, Aarsland D, Chaudhuri KR, Dobkin RD, Leentjens AF, Rodriguez-Violante M, et al. The neuropsychiatry of Parkinson’s disease: advances and challenges. Lancet Neurol 2022; 21: 89–102.

4 McKeith IG, Ferman TJ, Thomas AJ, Blanc F, Boeve BF, Fujishiro H, et al. Research criteria for the diagnosis of prodromal dementia with Lewy bodies. Neurology 2020; 94: 743–55.

5 Berg D, Borghammer P, Fereshtehnejad S-M, Heinzel S, Horsager J, Schaeffer E, et al. Prodromal Parkinson disease subtypes — key to understanding heterogeneity. Nat Rev Neurol 2021; 17: 349–61.

6 McKeith IG, Boeve BF, Dickson DW, Halliday G, Taylor J-P, Weintraub D, et al. Diagnosis and management of dementia with Lewy bodies. 2017.

7 Svendsboe E, Terum T, Testad I, Aarsland D, Ulstein I, Corbett A, et al. Caregiver burden in family carers of people with dementia with Lewy bodies and Alzheimer’s disease. Int J Geriatr Psychiatry 2016; 31: 1075–83.

8 Kanemoto H, Sato S, Satake Y, Koizumi F, Taomoto D, Kanda A, et al. Impact of Behavioral and Psychological Symptoms on Caregiver Burden in Patients With Dementia With Lewy Bodies. Front Psychiatry 2021; 12: 753864.

9 Wetmore JB, Li S, Yan H, Irfan M, Rashid N, Peng Y, et al. Increases in institutionalization, healthcare resource utilization, and mortality risk associated with Parkinson disease psychosis: Retrospective cohort study. Parkinsonism Relat Disord 2019; 68: 95–101.

10 Badenoch JB, Paris A, Jacobs BM, Noyce AJ, Marshall CR, Waters S. Neuroanatomical and prognostic associations of depression in Parkinson’s disease. J Neurol Neurosurg Psychiatry 2024;: jnnp-2023-333007.

11 Cummings J, Lanctot K, Grossberg G, Ballard C. Progress in Pharmacologic Management of Neuropsychiatric Syndromes in Neurodegenerative Disorders: A Review. JAMA Neurol 2024; 81: 645.

12 Fujishiro H, Iwata-Endo K, Kobayashi R, Morikawa F, Ikeda M. Electroconvulsive therapy for dementia with Lewy bodies: A systematic review and Japanese multicenter survey. Asian J Psychiatry 2025; 108: 104510.

13 Kazui H, Yoshiyama K, Kanemoto H, Suzuki Y, Sato S, Hashimoto M, et al. Differences of Behavioral and Psychological Symptoms of Dementia in Disease Severity in Four Major Dementias. PLOS ONE 2016; 11: e0161092.

14 Jaimes-Albornoz W, Ruiz De Pellon-Santamaria A, Nizama-Vía A, Isetta M, Albajar I, Serra-Mestres J. Catatonia in older adults: A systematic review. World J Psychiatry 2022; 12: 348–67.

15 Aarsland D, Perry R, Larsen JP, McKeith IG, O’Brien JT, Perry EK, et al. Neuroleptic Sensitivity in Parkinson’s Disease and Parkinsonian Dementias. J Clin Psychiatry 2005; 66: 504–14.

16 Kurlan R, Cummings J, Raman R, Thal L. Quetiapine for agitation or psychosis in patients with dementia and parkinsonism. Neurology 2007; 68: 1356–63.

17 Postuma RB, Berg D, Stern M, Poewe W, Olanow CW, Oertel W, et al. MDS clinical diagnostic criteria for Parkinson’s disease. Mov Disord 2015; 30: 1591–601.

18 Emre M, Aarsland D, Brown R, Burn DJ, Duyckaerts C, Mizuno Y, et al. Clinical diagnostic criteria for dementia associated with Parkinson’s disease. Mov Disord 2007; 22: 1689–707.

19 Benamer HTS, Oertel WH, Patterson J, Hadley DM, Pogarell O, Höffken H, et al. Prospective study of presynaptic dopaminergic imaging in patients with mild parkinsonism and tremor disorders: Part 1. Baseline and 3-month observations. Mov Disord 2003; 18: 977–84.

20 Chahid Y, Sheikh ZH, Mitropoulos M, Booij J. A systematic review of the potential effects of medications and drugs of abuse on dopamine transporter imaging using [123I]I-FP-CIT SPECT in routine practice. Eur J Nucl Med Mol Imaging 2023; 50: 1974–87.

21 Komatsu J, Samuraki M, Nakajima K, Arai H, Arai H, Arai T, et al. ^123^ I-MIBG myocardial scintigraphy for the diagnosis of DLB: a multicentre 3-year follow-up study. J Neurol Neurosurg Psychiatry 2018; 89: 1167–73.

22 Stefanelli A, Treglia G, Bruno I, Rufini V, Giordano A. Pharmacological interference with 123I-metaiodobenzylguanidine: a limitation to developing cardiac innervation imaging in clinical practice?.

23 Busner J, Targum SD. The clinical global impressions scale: applying a research tool in clinical practice. Psychiatry Edgmont Pa Townsh 2007; 4: 28–37.

24 Morikawa F, Kobayashi R, Murayama T, Fukuya S, Tabata K, Fujishiro H, et al. Evaluating Electroconvulsive Therapy for Dementia With Lewy Bodies, Including the Prodromal Stage: A Retrospective Study on Safety and Efficacy. Int J Geriatr Psychiatry 2024; 39: e70020.

25 Grover S, Sahoo S, Rabha A, Koirala R. ECT in schizophrenia: a review of the evidence. Acta Neuropsychiatr 2019; 31: 115–27.

26 Rohland BM, Carroll BT, Jacoby RG. ECT in the treatment of the catatonic syndrome. J Affect Disord 1993; 29: 255–61.

27 Van Der Does Y, Turner RJ, Bartels MJH, Hagoort K, Metselaar A, Scheepers F, et al. Outcome prediction of electroconvulsive therapy for depression. Psychiatry Res 2023; 326: 115328.

28 Leong IL, Ng TH, Sen K, Burchill E, Costello H, Badenoch JB, et al. Reduced striatal dopamine transmission as a transdiagnostic substrate of psychomotor retardation. Brain 2025;: awaf335.

29 Jauhar S, McCutcheon RA, Nour MM, Veronese M, Rogdaki M, Bonoldi I, et al. Dopamine and Mood in Psychotic Disorders: An^18^ F-DOPA PET Study. JAMA Psychiatry 2025; 82: 1009.

30 Landau AM, Chakravarty MM, Clark CM, Zis AP, Doudet DJ. Electroconvulsive Therapy Alters Dopamine Signaling in the Striatum of Non-human Primates. Neuropsychopharmacology 2011; 36: 511–8.

31 Kobayashi K, Imoto Y, Yamamoto F, Kawasaki M, Ueno M, Segi-Nishida E, et al. Rapid and lasting enhancement of dopaminergic modulation at the hippocampal mossy fiber synapse by electroconvulsive treatment. J Neurophysiol 2017; 117: 284–9.

32 Jellinger KA. Depression in dementia with Lewy bodies: a critical update. J Neural Transm 2023; 130: 1207–18.

33 Kazmi H, Walker Z, Booij J, Khan F, Shah S, Sudre CH, et al. Late onset depression: dopaminergic deficit and clinical features of prodromal Parkinson’s disease: a cross-sectional study. J Neurol Neurosurg Psychiatry 2021; 92: 158–64.

34 Nishida H, Takamiya A, Kudo S, Uchida T, Yamagata B, Bun S, et al. Characteristics of Severe Late-Life Depression in the Prodromal Phase of Neurodegenerative Dementia. Am J Geriatr Psychiatry Open Sci Educ Pract 2025; 5: 10–20.

